# Evaluation of the ROX index in SARS-CoV-2 Acute Respiratory failure treated with both High-Flow Nasal Oxygen (HFNO) and Continuous Positive Airway Pressure (CPAP)

**DOI:** 10.1101/2021.03.23.21254203

**Authors:** Hakim Ghani, Michael Shaw, Phyoe Pyae, Rigers Cama, Meghna Prabhakar, Alessio Navarra, Rahul Mogal, Andrew Barlow, Nazril Nordin, Rama Vancheeswaran, Janice Yu Ji Lee, Felix Chua

**Affiliations:** Respiratory Department, West Herts NHS Trust, Watford, UK; Intensive Care, West Herts NHS Trust, Watford, UK; Radiology Department, Addenbrookes Hospital, Cambridge, UK; Respiratory Department, Royal Brompton and Harefield NHS Trust, London, UK

**Keywords:** Acute Respiratory Failure, Continuous Positive Airway Pressure, High-Flow Nasal Oxygen, ROX index, SARS Coronavirus-2

## Abstract

**Background:** Non-invasive respiratory support including high-flow nasal oxygen (HFNO), and continuous positive airway pressure (CPAP) have been used to provide therapy in selected SARS-CoV-2 patients with acute respiratory failure (ARF). The value of the ROX index, a validated benchmark for outcomes in HFNO is unknown in CPAP.

**Objective:** Can the ROX, a validated benchmark in HFNO be used for measuring treatment outcomes of CPAP in SARS-COV-2 ARF?

**Study Design and Methods:** A non-randomised prospective protocol driven observational non-intensive care unit study in 130 SARS-COV-2 patients with ARF treated with non-invasive therapy from March 2020 to January 2021. The primary end point was failure of therapy (death or escalation). Secondary outcomes included time to failure including invasive mechanical ventilation (IMV) or death, the effect of escalation to CPAP from HFNO and the utility of ROX in ARF.

**Results:** HFNO was better than CPAP in treating SARS-COV-2 ARF: 17/35 (48.5%) with successful HFNO therapy versus 24/95 (25.2%) with CPAP. The ROX index was more sensitive to outcomes with CPAP compared to HFNO and distinguished treatment failure early at 1, 4, 6, 12, and 24 hours with the highest sensitivity at 24 hours (ROX-24h). The AUC for the ROX-24h was 0.77 for HFNO (P<0.0001), and 0.84 for CPAP (P<0.0001). The ROX-24h cut-points predicted failure with HFNO when < 3.9 (PPV 71%, NPV 75%) and CPAP < 4.3 (PPV 75%, NPV 91%). For success, ROX-24h cut-points of 7.6 for HFNO (PPV 85%, NPV 48%) and 6.1 for CPAP (PPV 88%, NPV 62%) were observed. Escalation from HFNO to CPAP was mostly not successful.

**Conclusion:** ARF in SARS-COV-2 can be successfully managed by non-invasive support. The ROX index, validated for HFNO, provides a timely, low resource measure for both HFNO and CPAP avoiding delayed intubation.

**Trial registration:** Study approved by NHS HRAREC (20/HRA/2344;ethics 283888)

**KEY MESSAGE:** *What is the key question?:* Can the ROX, a validated benchmark in high-flow nasal oxygen (HFNO) be used for measuring treatment outcomes of continuous positive airway pressure (CPAP) in SARS-COV-2 ARF?

*What is the bottom line?:* The ROX index, validated for HFNO, provides a timely, low resource measure for both HFNO and CPAP support avoiding delayed intubation.

*Why read on?:* The present study compares the efficacy of HFNO and CPAP, two common globally used modalities of treatment for SARS-CoV-2 and notes the superior utility of the ROX-24h in CPAP to predict outcome, enabling timely escalation decisions.

## INTRODUCTION

The overwhelming burden of patients with hypoxic respiratory failure due to SARS coronavirus-2 (SARS-COV-2) on intensive care units (ICUs) is well documented ^1–5^. Comparison of outcomes across countries is difficult because of data heterogeneity arising from variations in experience, bias, resource, concerns about aerosols and the absence of standardized measurements for both therapies^6–8^. First-wave experience in Italy, needed an expanded ICU capacity that administered 11% non-invasive ventilation (NIV) (continuous positive airway pressure, CPAP) in addition to 88% invasive mechanical ventilation (IMV)^1^. The French REVA investigators used high-flow nasal oxygen (HFNO) in 20% and 6% CPAP, reducing IMV to 63%^2^. Mortality was higher in patients treated with IMV when intubated early (within 24 hours) or after 5 days^3,9^. Early reports note a higher proportion of hospitalised patients with SARS-COV-2 ARF were intubated within 24 hours with higher mortality^,3,9^. This risk appears attenuated by HFNO and CPAP where a review of 21 non-randomised studies reported success rates between 55 – 60% for each arm^6,8^.

Most countries have surged into respiratory wards due to demand from SARS-CoV-2 ARF^9–13^ where non-invasive respiratory support has traditionally been administered^2,3,13,14^. Trials are in progress to compare non-invasive therapies: HFNO, and CPAP to maximise outcomes^15,16^. Existent studies do not reliably provide comparison as they are mostly single arm with differing baseline characteristics, particularly immune-suppression, an important modifying factor affecting ventilatory outcomes^1,8,13,17,18^. The optimal non-invasive treatment, if any, for managing ARF in SARS-COV-2 is not known.

Current guidelines are broad in scope but conflicting. Prevailing British NICE guidance is to avoid HFNO whereas the British Thoracic Society recommends either, based on HFNO reports^19,20^. The World Health Organisation interim guidance suggested selected use of NIV in SARS-COV-2^6^ while the Surviving Sepsis Guideline recommends HFNO in preference to CPAP^21^. A recent European algorithm recommends HFNO escalation to CPAP, prior to IMV^22^ conflicting with reports where this delays intubation worsening mortality^6,23^. Recovery-Respiratory Support, a UK trial, aims to address this equipoise but excludes patients escalated from HFNO to CPAP^15,22^and may highlight national bias.

A benchmark for success of non-invasive respiratory support will enable easier comparison but has yet to emerge^24,25^. The ROX index, a ratio of oxygenation and respiratory rate, has been validated in SARS-CoV-2 for HFNO^10,11,13,14,26^. It’ s use in CPAP is reported in small studies with conflicting results^11,27^. In this study, we report the superior efficacy of HFNO compared with CPAP in the treatment of SARS-COV-2 ARF and importantly note the utility of the ROX, particularly in CPAP as an early benchmark of therapy outcome.

## METHODS

### Study design and baseline characteristics

We conducted a prospective single site non-randomised protocolised observational study in SARS-CoV-2 ARF in a large UK district hospital with over 2500 hospital assessments over 12 months. Adults older than 18 years who tested positive for SARS-CoV-2 nucleic acid by real-time reverse transcriptase polymerase chain reaction (rRT-PCR) and ARF were analysed retrospectively, where a full prospectively collected dataset was available. This was part of a National Health Service Health Research Authority (20/HRA/2344; ethics reference: 283888) approved study which included patient and public participation in the observational design. Baseline clinical characteristics and investigation results were collected according to a pre-specified protocol^28^.

### Laboratory, physiologic and radiographic data

Laboratory tests, radiographs, and physiological measurements were performed as part of routine clinical care. Nasopharyngeal swabs for rRT-PCR were tested in UK Public Health England laboratories. Baseline observations included all the parameters recommended by the National Early Warning Score. The ROX index defined as the ratio of peripheral oxygen saturations (oximetry, SpO2) to the product of respiratory rate and fraction of inspired oxygen (FiO2) was calculated from observations^29^. Chest radiographs acquired in ED were assessed by 2 radiologists: each lung field was divided into upper, middle, and lower zones and scored 1 point for each zone affected. If performed during the admission, the first CT scan was scored by assessing each lobe for inflammatory changes using the semi-quantitative method of Francone et al^30^; briefly, 0-5 points were assigned per lobe based on visually assessed involvement: 0 points for none; 1 point for <5%; 2 points for 5-25%; 3 points for 26-49%; 4 points for 50-75%; and 5 points for >75%. Total CT severity score was taken as the sum points given for each lobe (maximum of 25 points).

### Location and level of care

Following presentation, ARF was defined as a respiratory rate of >30/min with oximetry of <92% despite oxygen at 15litres/min via reservoir bag. The timing, type, and location of ARF support was made at the discretion of the treating respiratory and ICU physician based on clinical severity and availability of equipment. Twice daily multi-disciplinary team discussions enabled smooth transfer and communication of deteriorating patients. Awake prone positioning was encouraged and reinforced by all staff. All data collected for the study was assessed prospectively and did not inform decision making.

### Equipment

Non-invasive respiratory support was provided in well ventilated isolation units where staff were provided full PPE. HFNO was delivered by the Airvo 2 (Fisher & Paykel Healthcare) at flows initiated at 60L/min. CPAP was provided by NIV machines (V60, Trilogy, Respironics) via face mask interfaces with median positive end-expiratory pressure (PEEP) of 8 (6-12). FiO2 in both groups was titrated to target saturations of between 92-96%.

### Outcomes

The primary endpoints were failure which was considered when (1) death occurred during the episode, (2) escalation to another non-invasive modality or (3) mechanical ventilation (IMV) was required. Success was defined when patients improved on therapy and avoided death or escalation. Secondary outcomes were time to escalation and death. These were referenced to time of diagnosis and described as continuous lognormally distributed variables. Of secondary interest were predictors of failure, including baseline characteristics and measurement of the ROX at pre-specified timepoints.

### Statistical analysis

Descriptive statistics and comparisons between parametric and non-parametric samples were calculated using the GraphPad PRISM statistics software (GraphPad, San Diego, USA). Categorical variables were expressed as frequencies and percentages and compared using Pearson’ s⍰χ2 tests. Continuous variables were expressed as means + standard deviations (SDs) unless lognormal in distribution, where geometric means (GM) with geometric standard deviation (GSD) were presented. Normality and lognormality were assessed by the D’Agostino-Pearson Omnibus K2 test, review of QQ plots of residuals, and qualitative assessment of plausibility. Where continuous variables were identified as lognormally distributed, they were normalised by taking the natural logarithm of their values prior to further statistical analysis with parametric tests. Non-parametric data was compared using Wilcoxon rank-sum tests.

A CONSORT diagram reported the flow of patients (Figure 1). Kaplan-Meier curves were constructed to assess mortality between groups. Mantel-Cox Log-Rank tests were used to assess for difference in survival between groups (Figure 2A). Where treatment escalation from HFNO to CPAP was noted, the impact on gas exchange and time to intubation (IMV) (Figure 2B-C) was compared using paired t-tests. Correlations between the A-a gradient, PF (PaO2/FiO2) ratios, SF ratios and ROX scores was modelled by least-squares linear regression and analysed using the R2 coefficient of determination (Figure 3 A-D). ROX scores were calculated at various times using the FLORALI formula ^29^ for both HFNO and CPAP (Figure 4A and B). For each therapy, serial ROX scores were compared between outcome groups using two-way mixed effects models with Geisser-Greenhouse Sphericity Correction. Multiple comparison tests between group means at each time point were made with Šidàk’ s correction.

**Figure 1.**
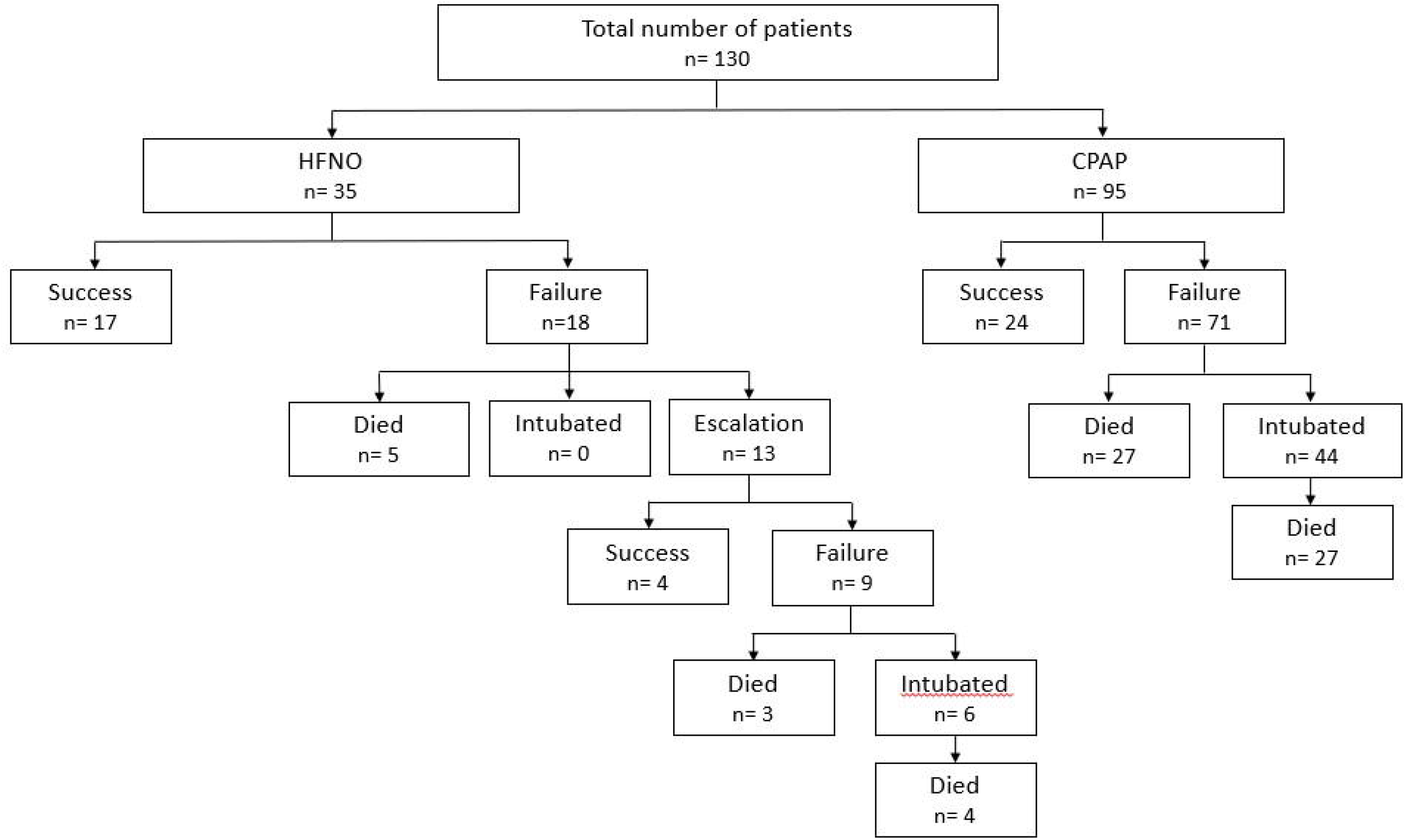
CONSORT Diagram

**Figure 2.**
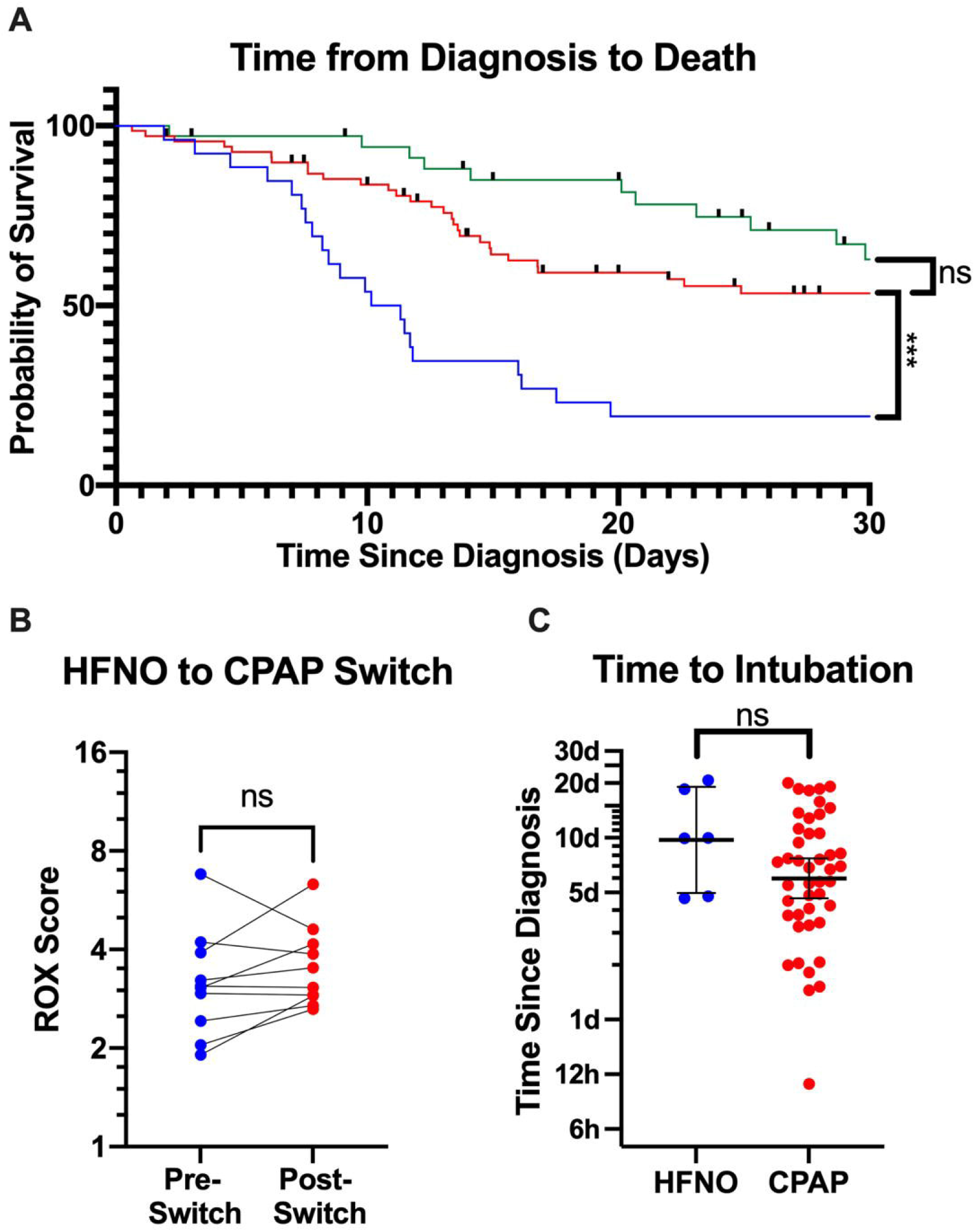
(2A) Kaplan Meier survival curves for patient treated with CPAP and HFNO in the first and second waves are shown. No Dexamethasone was used in Wave 1 CPAP patients. Figure 2B shows the change in ROX at 2 hours in patients who were started on CPAP after HFNO failure. No significant change was noted in the median ROX at 2 hours. Figure 2C shows time to IMV in those escalated from HFNO and CPAP.

**Figure 3.**
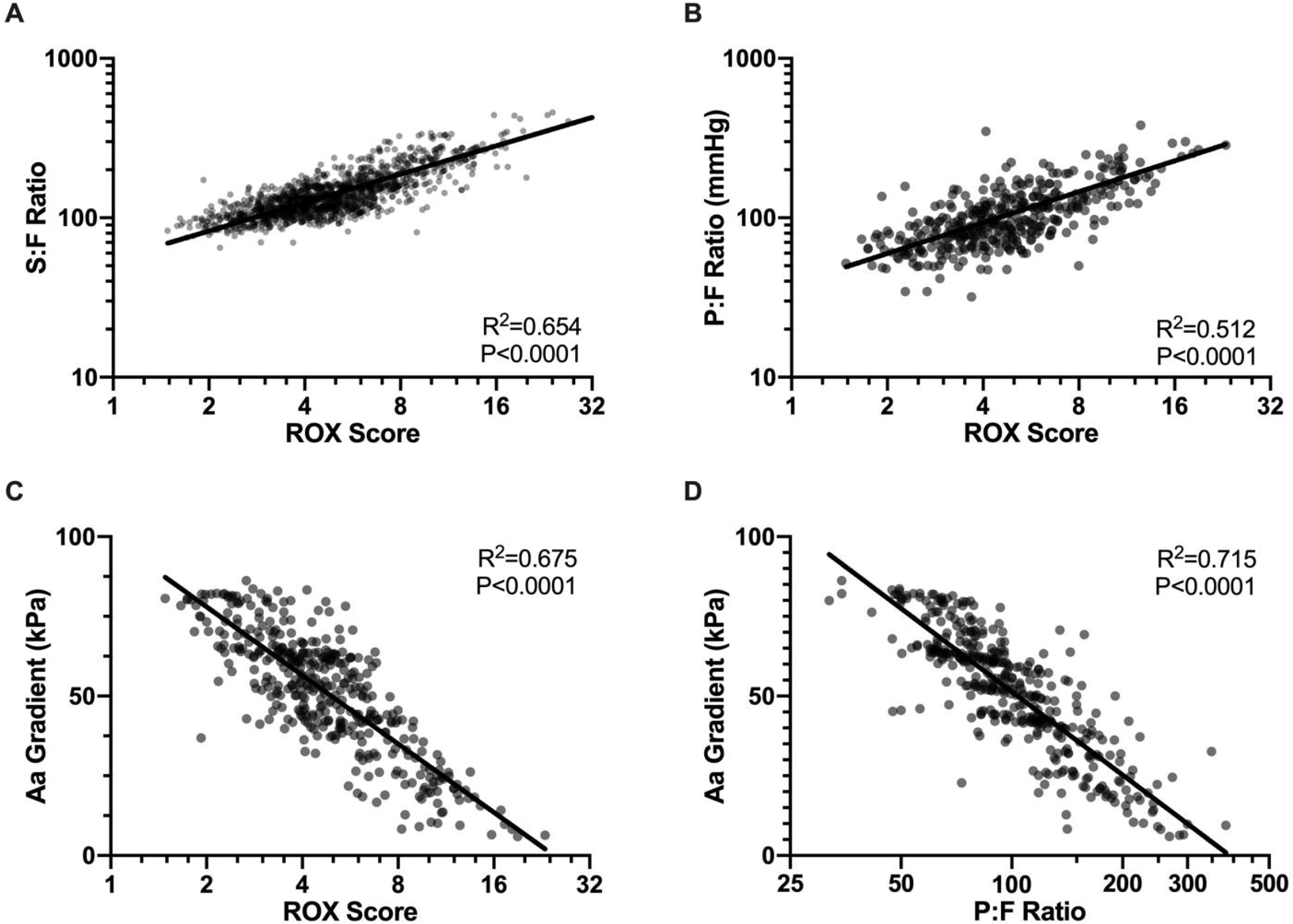
Figure 3a, b and c show the correlations between the ROX score, S:F ratio, P:F Ratio, and A-a gradient. (3A), S:F ratio and ROX. (3B) P:F ratio and ROX. (3C) A:a Gradient and ROX (3D) A:A Gradient and PF ratio. All correlations are significant with p<0.0001 for all).

**Figure 4.**
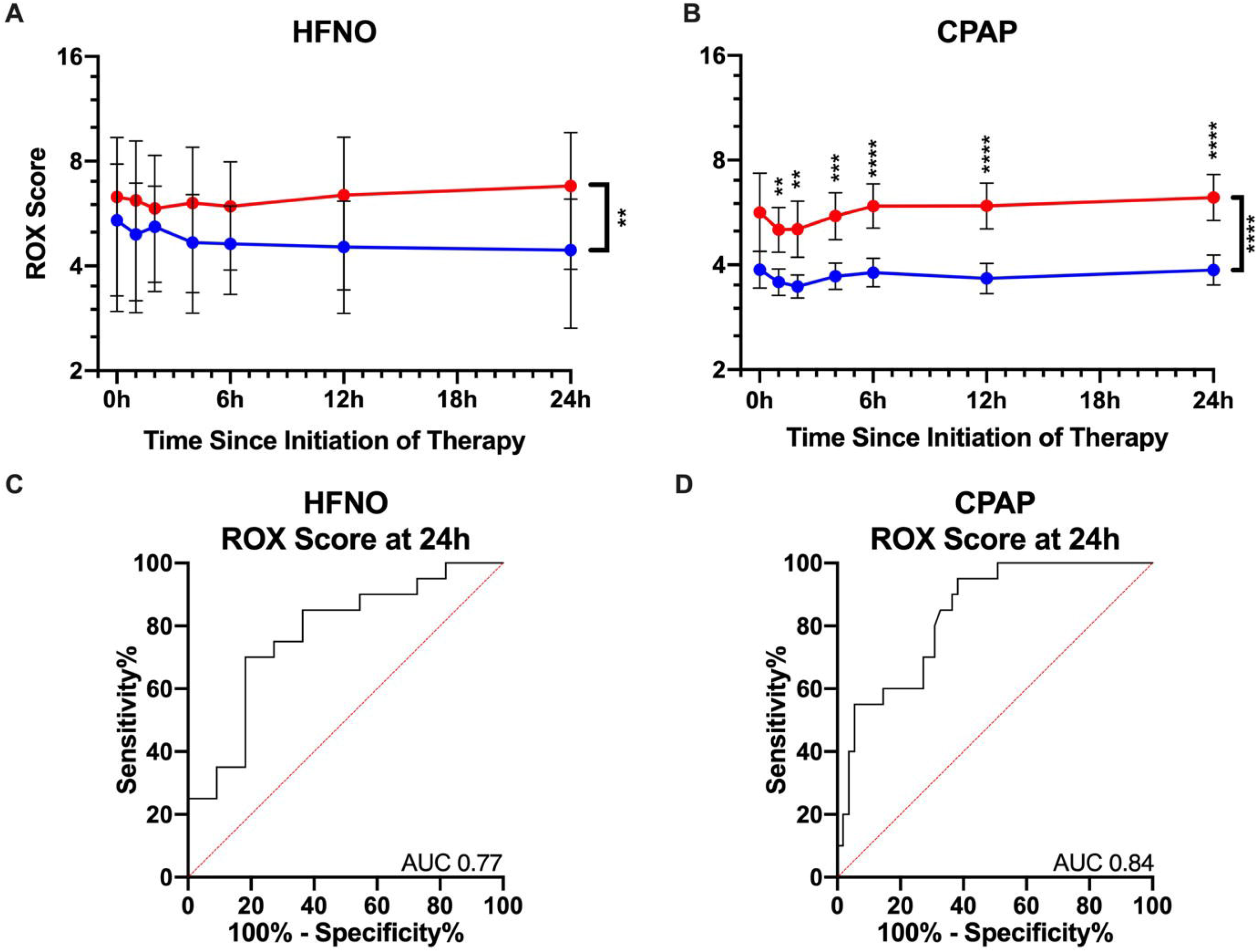
Figure 4A and 4B show the ROX scores over time in patients with failure (blue dots) or successful therapy (red dots). The Geometric mean and 95% CI are shown for patients who failed (blue lines) and who had successful outcomes (red lines) with p values. P<0.05, <0.005, p<0.0005 are denoted by **, ***, **** respectively. Figure 4C and 4D are the ROC curves for HFNO and CPAP at 24 hours

Receiver operating characteristic (ROC) curves were constructed using the ROX-24h post initiation of each therapy to assess their value in predicting case outcome (Figure 4C, 4D). Youden’ s index was calculated to determine ROX-24h cut-offs that maximised sensitivity and specificity for case outcomes. Negative predictive value (NPV) and positive predictive value (PPV) were estimated using published success rates of 60% and 55% for HFNO and CPAP respectively (Table 3 supplementary data).

## RESULTS

130 patients with ARF secondary to SARS-CoV-2 infection had a full data set that was analysed between March 2020 and January 2021: 35 were treated with HFNO (after UK restrictions were lifted in September 2020), and 95 were treated with face mask CPAP. Success was documented in 17/35 (48.5%) with HFNO and 24/95 (25.2%) with CPAP. In the first wave (March to July 2020), Dexamethasone and HFNO were not used. 26 patients in the first wave were treated with CPAP with a mortality of 21/26 (80.7%). In the second wave, where Dexamethasone was used in all patients, success was noted in 17/35 (48.5%) patients with HFNO compared with 21/69 (30.4%) with CPAP.

The baseline patient characteristics are presented in Table 1. The median age at presentation of patients with ARF was 60 years. Similar median age (IQR) in wave 1: 60 years (50-69) and wave 2: 59 years (46-72) was noted. The age distribution of patients was 30/130 (23%) in the <50-year-olds, 75/130 (58%) in the 50–70-year-olds and 25/130 (19%) in the over 70-year-olds. Baseline age did not differ between HFNO and CPAP outcomes (p=0.17).

**Table 1:** Baseline characteristics of patients and outcomes.

| Baseline Characteristics | Whole Cohort | HFNO |  | CPAP |  | p value |
| --- | --- | --- | --- | --- | --- | --- |
|  |  | Failure | Success | Failure | Success |  |
|  | N | n (%) | n (%) | n (%) | n (%) |  |
|  | 130 | 18 (14) | 17 (13) | 71 (55) | 24 (18) |  |
| Age | Mean ± SD (n) | Mean ± SD (n) | Mean ± SD (n) | Mean ± SD (n) | Mean ± SD (n) | |
| First wave | 60 ± 9.3 (26) | - | | 60 ± 9.5 (23) | 58 ± 8.5 (3) | 0.99<sup>a</sup> |
| Second Wave | 59 ± 13.5 (104) | 64 ± 11 (18) | 58 ± 11 (17) | 62 ± 13.1 (48) | 50 ± 13.5 (21) | 0.96<sup>c</sup> |
|  | n (%) | n (%) | n (%) | n (%) | n (%) |  |
| <50 | 30 (23) | 1 (6) | 6 (35) | 11 (15) | 12 (50) | 0.17<sup>b</sup> |
| 50-70 | 75 (58) | 13 (72) | 8 (47) | 44 (62) | 10 (42) |  |
| >70 | 25 (19) | 4 (22) | 3 (18) | 16 (23) | 2 (8) |  |
| Gender | n (%) | n (%) | n (%) | n (%) | n (%) |  |
| Male | 89 (68) | 13 (72) | 11 (65) | 50 (70) | 15 (63) | 0.99<sup>b</sup> |
| Smoking Status | n (%) | n (%) | n (%) | n (%) | n (%) |  |
| Current Smoker | 5 (4) | 1 (6) | 0 (0) | 4 (6) | 0 (0) | 0.77<sup>b</sup> |
| Ex-Smoker | 36 (28) | 5 (28) | 6 (35) | 15 (21) | 10 (42) |  |
| Never Smoker | 39 (30) | 7 (39) | 8 (47) | 16 (23) | 8 (33) |  |
| Not scored | 50 (38) | 5 (28) | 3 (18) | 36 (51) | 6 (25) |  |
| Comorbidities | n (%) | n (%) | n (%) | n (%) | n (%) |  |
| Obesity | 71 (55) | 13 (72) | 6 (35) | 40 (56) | 12 (50) | 0.90<sup>b</sup> |
| History of Stroke | 4 (3) | 2 (11) | 1 (6) | 1 (1) | 0 (0) | 0.87<sup>b</sup> |
| History of Dementia | 3 (2) | 1 (6) | 0 (0) | 2 (3) | 0 (0) | 0.99<sup>b</sup> |
| CKD Stage 1 | 25 (19) | 3 (17) | 3 (18) | 13 (18) | 6 (25) | 0.98<sup>b</sup> |
| CKD Stage 2 | 65 (50) | 9 (50) | 10 (59) | 32 (45) | 14 (58) |  |
| CKD Stage 3 | 35 (27) | 3 (17) | 4 (24) | 24 (34) | 4 (17) |  |
| CKD Stage 4 | 3 (2) | 1 (6) | 0 (0) | 2 (3) | 0 (0) |  |
| CKD Stage 5 | 1 (1) | 1 (6) | 0 (0) | 0 (0) | 0 (0) |  |
| Leucocytes > 11 | n (%) | n (%) | n (%) | n (%) | n (%) |  |
|  | 17 (13) | 5 (28) | 2 (12) | 9 (13) | 1 (4) | 0.90<sup>b</sup> |
| Lymphocytes < 0.7 | n (%) | n (%) | n (%) | n (%) | n (%) |  |
|  | 28 (22) | 4 (22) | 4 (24) | 13 (18) | 7 (29) | 0.99<sup>b</sup> |
| Baseline PF | Geometric mean * SD (n) | Geometric mean * SD (n) | Geometric mean * SD (n) | Geometric mean * SD (n) | Geometric mean * SD (n) |  |
| First Wave | 86.08 * 1.44 (25) | - | - | 81.65 * 1.40 (22) | 127.00 * 1.40 (3) | 0.90<sup>a</sup> |
| Second Wave | 106.8 * 1.69 (95) | 144.00 * 1.78 | 107.30 * 1.73 (17) | 94.38 * 1.66 (44) | 108.70 * 1.52 (19) | 0.57<sup>c</sup> |
| Rox | Geometric mean * SD (n) | Geometric mean * SD (n) | Geometric mean * SD (n) | Geometric mean * SD (n) | Geometric mean * SD (n) |  |
| First Wave | 2.89 * 1.32 | - | - | 2.77 * 1.29 | 3.97 * 1.22 | 0.00002 |
|  |  |  |  | (23) | (3) | <sup>a</sup> |
| Second Wave | 5.45 * 1.81 | 6.89 * 1.93 | 6.00 * 1.67 (17) | 4.43 * 1.68 (48) | 6.37 * 1.94 (21) | 0.48 <sup>c</sup> |
| SOARS Score | Median (Q1-Q3) | Median (Q1-Q3) | Median (Q1-Q3) | Median (Q1-Q3) | Median (Q1-Q3) |  |
|  | 6 (5-8) | 7 (5-9) | 7 (5-8) | 6 (5-8) | 5.5 (5-7) | 0.90 <sup>b</sup> |
| CXR affected zones | Median (Q1-Q3) | Median (Q1-Q3) | Median (Q1-Q3) | Median (Q1-Q3) | Median (Q1-Q3) |  |
|  | 4 (2-5) | 2.5 (0.75-4) | 4 (3.5-5) | 4 (2-5) | 4 (3-5) | 0.87 <sup>b</sup> |
| CT Score | Median (Q1-Q3) | Median (Q1-Q3) | Median (Q1-Q3) | Median (Q1-Q3) | Median (Q1-Q3) |  |
|  | 23 (20.5-25) | 25 (21-25) | 24 (21.25-25) | 23.5 (13.75-25) | 22.5 (19-23.25) | 0.99 <sup>b</sup> |
| Dexamethasone | n (%) | n (%) | n (%) | n (%) | n (%) |  |
|  | 103 (79) | 18 (100) | 17 (100) | 48 (68) | 20 (83) | 0.03 <sup>b</sup> |
| Invasive support (days) | Geometric mean * SD | Geometric mean * SD | Geometric mean * SD | Geometric mean * SD | Geometric mean * SD |  |
|  | 2.62 * 4.32 | 3.90 * 3.93 | 3.43 * 3.03 | 2.44 * 4.79 | 2.30 * 4.35 | 0.99 <sup>b</sup> |
| Time to IMV (days) | Geometric mean * SD | Geometric mean * SD | Geometric mean * SD | Geometric mean * SD | Geometric mean * SD |  |
|  | 6.34 * 2.27 | 9.73 * 1.9 | - | 5.99 * 2.29 | - | 0.87 <sup>b</sup> |
| Mortality | n (%) | n (%) | n (%) | n (%) | n (%) |  |
|  | 66 (51) | 12 (67) | - | 54 (76) | - | 0.99 <sup>b</sup> |

a: difference between waves
b: difference between groups
c: baseline second wave

Males were similarly distributed between HFNO and CPAP with no difference in outcomes: HFNO, 24/35 (68%), CPAP 65/95 (68%), HFNO or CPAP outcomes (p=0.99). Obesity defined as BMI>30, was noted in 71/130 (55%) patients with similar distribution in HFNO 19/35 (54%) and CPAP 52/95 (54.7%), p=0.90. Obesity did not influence HFNO failure 13/18 (72%) compared with CPAP failure (40/71, 56%) (p=not significant, ns). A smoking history was noted in 12/35 (34%) HFNO patients and 29/95 (30%) CPAP patients, with no significant difference between outcomes, p=0.77. Comorbidities including stroke, dementia, chronic kidney disease were not different between the arms or with different outcomes (see table 1). Inflammatory markers including leucocytosis and lymphopenia, selected as relevant in multivariate assessment of risk in COVID 19 ^28^ were not different at baseline or between therapy outcomes: 7/35 (20%) HFNO patients and 10/95 (10.5%) CPAP patients had a raised WCC and 8/35 HFNO (22%) and 20/95 CPAP patients (21%) had a low lymphocyte count. Radiological severity assessed by CXR zonal scores were a median of 2.5/6 in HFNO failure and 4/6 in CPAP failure, p=0.87 (ns). CT scores were a median of 25/30 in HFNO failure and 23.5/30 in CPAP failure with no significant difference between HFNO and CPAP or with outcomes (p=0.99, ns).

The baseline PaO2/FiO2 ratio (PF) for the whole cohort was 86.08 in the first wave and 106.8 in the second wave which was not significantly different (p= 0.90). This was different to the baseline ROX score between wave 1 and wave 2 where wave 1 was more severe with a geometric mean of 2.89 ± 1.44 compared with 5.45 ± 1.81, p=0.00002. The ROX was compared between patients in wave 2 for HFNO and CPAP outcomes. In wave 2, the mean ROX was similar at baseline between HFNO and CPAP patients and between HFNO success (6.00) and CPAP success (6.37). The baseline ROX scores in those with CPAP failure were lower (4.43+1.68) than HFNO failure (6.89 + 1.93) but did not reach statistical significance p=0.48.

There was a clear difference in CPAP outcomes between the first and second waves where dexamethasone was used variably. Dexamethasone was given to all HFNO patients as they were treated after the RECOVERY trial^17^ results: 35/35 (100%) compared with 69/95 (72%) patients in the CPAP arm. Failure with Dexamethasone and HFNO was 18/35, 51% compared with Dexamethasone and CPAP 48/69 (70%).

Success with HFNO was noted in 17/35 patients (50%) and in 24/95 (25%) with CPAP (Wave 1: 12%, Wave 2: 30%). HFNO failure was escalated to CPAP in 13 patients with success in 4/13 (31%). 5/18 patients had a ceiling of care decision for ward-based care where no further escalation was considered. No patients were intubated without a trial of CPAP. Mortality in the intubated HFNO group was 4/6 (HFNO-IMV, 67%). Mortality during the hospital episode for all HFNO patients 12/35 (31%). 22/35 (63%) patients were treated on the ward. In the CPAP group, 24/95 (26%) were successfully treated while 71/95 failed (75%), 27/95 (28%) had a palliative management plan and died on the ward. 44/71 (62%) were intubated, of which 27/44 died (CPAP-IMV, 61%). The time to start of respiratory support was not significantly different in the HFNO group compared with CPAP (3.3d ±3.5 vs 2.35d ± 4.5, p=0.99). Time to intubation in the HFNO group was longer than those in the CPAP group but was not significant (9.73d ± 1.9 vs 5.99d ± 2.99, p= 0.87). This was due to escalation with an interim trial of CPAP prior to IMV, where delayed intubation was noted of 3 days.

Figures 3A-D show the ROX score correlations with the SF ratio, PF ratio and A-a gradient. Close relationships were reported with all 3 but highest with the A-a gradient (r^2^= 0.675, p<0.0001), less with the PF ratio (r^2^=0.512, p<0.0001) and SF ratio (r^2^=0.654, p<0.0001). The A-a gradient did correlate well with the PF ratio (r^2^=0.715, p<0.0001).

Figure 4A and B show the hourly change in ROX scores for patients treated with HFNO and CPAP where comparable measurement ranges are noted (ROX scores between 3-7) with divergent trends between success and failure of therapy for HFNO but more parallel trends with CPAP. A significant result was noted only for the pooled data for HFNO outcomes (p=0.0044) with a trend showing divergence between patients at 4, 6,12 and 24 hours (p=ns at all discrete time points). Those treated with CPAP showed no baseline difference but with significant parallel separation at 1,2,4, 6, 12, and 24 hours. (The Table 2 supplementary data shows the median ROX scores at all time points). The AUC for HFNO and CPAP at 24h (ROX-24h) are shown in Figures 4C and 4D. The ROX-24h AUC for HFNO was 0.77, p<0.0001 and ROX-24h for CPAP was 0.84, p<0.0001. The ROX-24h cut-point predicting high likelihood for failure with HFNO was 4.19 and CPAP was 4.29 (Table 3 supplementary data with sensitivity and specificity). A positive predictive value for failure with HFNO at 24 hours is 3.9 (PPV 71%, NPV 75%) and CPAP 4.3 (PPV 75%, NPV 91%). For success, cut points of 7.6 for HFNO (PPV 85%, NPV 48%) and 6.1 for CPAP (PPV 88%, NPV 62%) are noted.

## DISCUSSIONS

This prospective non-randomised protocolised single site observational study compared two commonly used non-invasive respiratory modalities: HFNO and CPAP in SARS-COV-2 ARF and evaluated the ROX index as an early benchmark in both arms, in a real-life pandemic setting. We report better outcomes with HFNO including mortality, escalation, intubation rates (IMV) compared with CPAP with a confirmatory ROX index value similar to prior studies^,10,11,13,14^. More significantly, we report sensitivity of the ROX index in CPAP which indicates likely success from 2 hours with maximal sensitivity at 24 hours, confirming early signals from smaller studes^25,27^. HFNO and CPAP are used globally to manage the demand posed by the SARS-CoV-2 pandemic. The utility of the ROX in predicting outcome within 24 hours should enable decisive escalation in the appropriate window of opportunity for IMV, reducing mortality (24 hours to 5 days)^7,3,9,13^.

The alveolar arterial gradient is the gold standard for measurement of severity in ARF^31^. The components of the A-a gradient are measures of ventilation-perfusion (VQ) mismatch, diffusion abnormalities and changes in ventilation (Co2). In ARF due to SARS-COV-2, patients present with severe hypoxemic hypocapnoeic alkalosis where it has been speculated that the PaO2/FiO2 (PF) ratio is an underestimate of hypoxaemia unlike the A-a gradient, because the PaO2 values standardised to PaCO2 levels is not calculated (1,66*PaCO2-PaO2 – 66,4 mmHg)^12,23^. We show a higher correlation between the ROX index and the A-a gradient, as opposed to the PF ratio. The practical ease of measurement of the ROX, in contrast to arterial blood sampling makes it easy for repetitive measurements using existent observations. Previous investigators have suggested the use of the SF (SpO2/FiO2) ratio^12^ which may be prone to inaccuracies of the oxygen dissociation curve with saturations < 92%. This limitation has been reported to be mitigated by the addition of the respiratory rate: the ROX index^23^.

We document improved mortality with HFNO compared with face mask CPAP like numerous others^10,13,14,21,26,27^. The ranges of measurements for the ROX were similar for HFNO and CPAP in SARS-CoV-2 ARF. For HFNO, we note a similar ROX at 4 hours to the French investigators (>5.37 for success) ^10,14^ and UK reports at 12 hours (ROX-12h <4.88 for failure)^11^. We note different results to a South African study (<3.7 for failure versus <4.46)^13^ which may be explained by the younger age of the population (52y vs 60y), a lower presenting PF ratio (68 vs 106) and differing treatment regimens (70% corticosteroid use) and comorbidities particularly HIV. The increased sensitivity of the ROX with time is well reported particularly at or before 24 hours aiming to maximise decisions about escalation early^11,13^. While our HFNO data is not significant at 4 and 6 hours, the trend with time is clearly significant with divergence at 4 hours and may reflect the smaller HFNO patient numbers in our study, a limitation due to the late UK adoption of HFNO.

We provide an early positive report of the value of the ROX in CPAP, unlike the small studies in the past with 20 and 18 patients who report mixed results^11,25^. The ROX discriminates outcomes early from 1 hour onwards with the most sensitivity at 24 hours. This should enable timely decision making, particularly in the ‘golden window’ between 24 hours and before 5 days^3,9^. No added benefits of CPAP is provided to those with a high BMI, a perception that often biases treatment choice and may account for the sicker baseline ROX in the CPAP failure group^32^. A practice in the UK and Europe is the escalation from HFNO to CPAP due to perception of superiority from additional PEEP^6,32^. Reports of PEEP induced injury, less tolerance, delayed intubation confer risk to this untested practice^6,21,29^. The ROX-24h provides reliable, time sensitive information that predicts outcome and provides a comparator for HFNO and CPAP allowing objective early indication of failure needing IMV. Our study shows that the ROX score did not consistently improve in most patients escalated from HFNO to CPAP. A responder analysis did show that 25% of patients improved avoiding IMV and that the ROX indicated those likely to benefit by 2 hours. While current trials are comparing HFNO, CPAP and standard oxygen in ARF due to SARS-COV-2, patients escalated between the arms are excluded and may not provide data on this untested but common practice^15^.

The many limitations of this study include the single-site, non-randomised observational design which may introduce unconscious bias in the choice of therapy and account for the lower baseline ROX in our CPAP failures. Secondly, the smaller HFNO group and the latter introduction of Dexamethasone adds variability but reflects the real-life pandemic UK practice with evolving guidance. Lastly, infectious risk was not measured as this has been extensively reported and was mitigated with appropriate PPE in well ventilated isolation units^7^.

We provide an initial report of the ROX index in a large group of patients treated with CPAP for SARS-CoV-2 ARF. Notably, we report that the ROX-24h is a timely benchmark of therapeutic efficacy in patients treated either CPAP or HFNO enabling decisive management at 24 hours. Prospective studies of the ROX in multi-site studies is indicated to explore its impact on mortality.

## Data Availability

All data is made available on the manuscript

## 1. Guarantor statement

Rama Vancheeswaran is the guarantor of the content of the manuscript, including the data and analysis

## 2. Author contributions

RV, HG, NN: Study design, study supervision, literature search, data collection, data interpretation, writing/revision of manuscript.

MS, PP, RC, MP, AN, JL: data collection, data analysis, revision of manuscript

FC, RM, MP, AB: study design, data interpretation, revision of manuscript

---

**Table 1 (Supplementary data):**
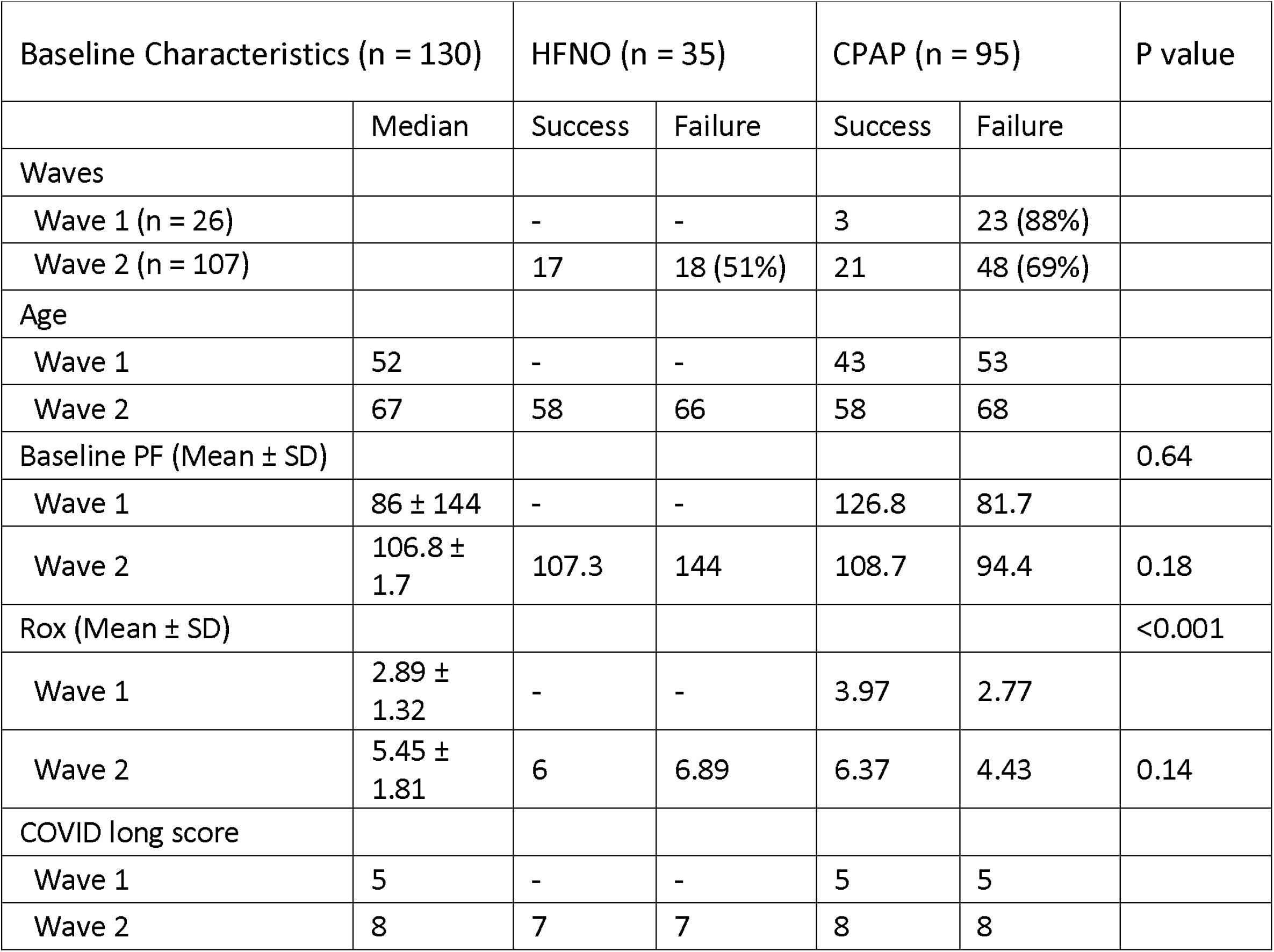
Baseline characteristics of patients in the 2 waves and outcomes.

**Table 2 (Supplementary data):**
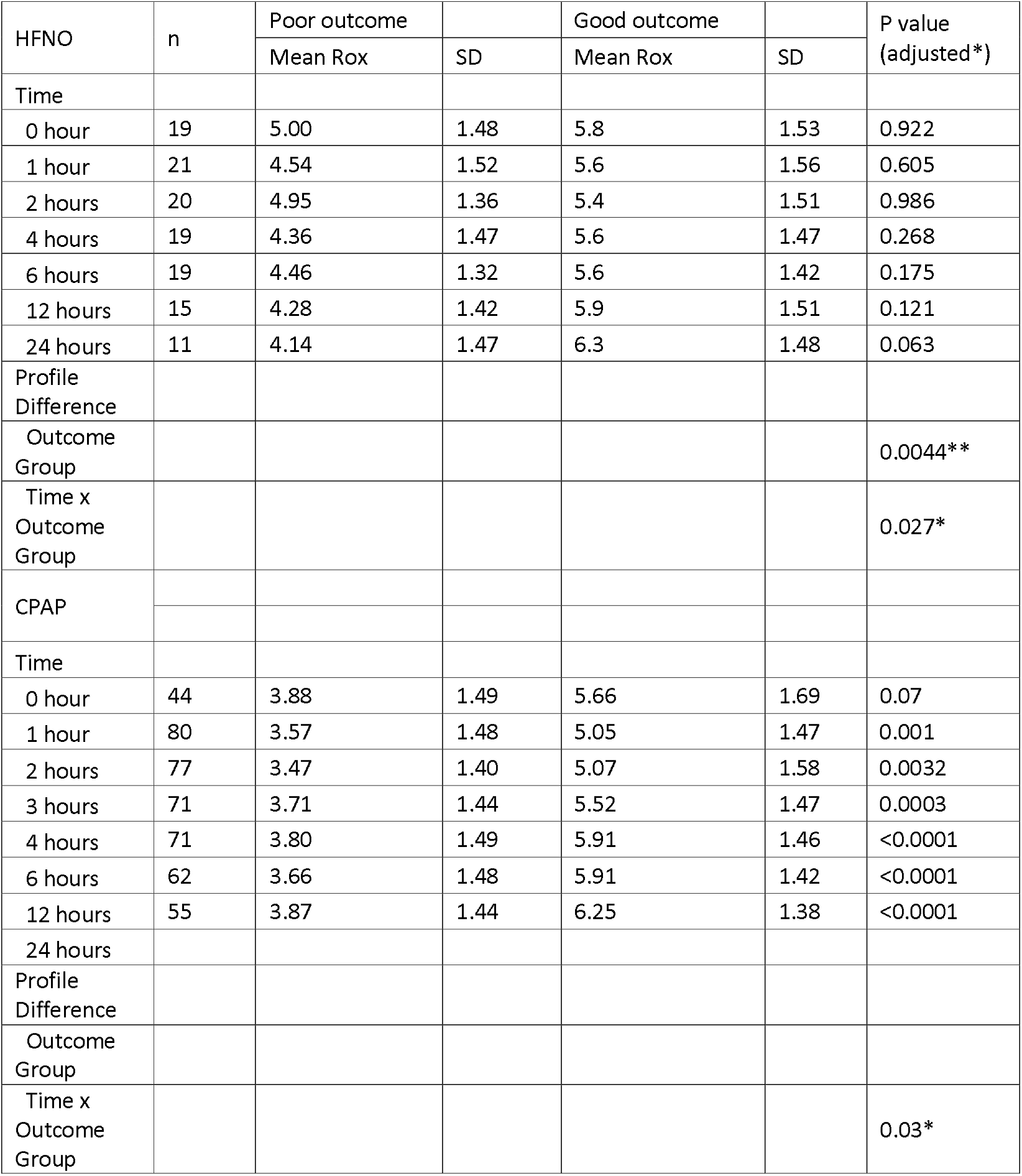
Differences between HFNO and CPAP over time.

**Table 3 (Supplementary data):** Rox AUC sensitivity and specificity for HFNO and CPAP at 24 hours.

| HFNO ROX-24 |  |  |  |  |  |  |  |
| --- | --- | --- | --- | --- | --- | --- | --- |
| Cut-point | Sensitivity | 95% CI | Specificity | 95% CI | Youden | PPV | NPV |
| > 3.926 | 90 | 69.90% to 98.22% | 45.5 | 21.27% to 71.99% | 35.35 | 71.20% | 72.50% |
| > 4.159 | 85 | 63.96% to 94.76% | 63.6 | 35.38% to 84.83% | 48.64 | 77.80% | 73.90% |
| > 7.648 | 35 | 18.12% to 56.71% | 90.9 | 62.26% to 99.53% | 25.91 | 85.20% | 48.30% |
| CPAP ROX-24 |  |  |  |  |  |  |  |
| Cut-point | Sensitivity | 95% CI | Specificity | 95% CI | Youden | PPV | NPV |
| > 4.290 | 95 | 76.39% to 99.74% | 61.8 | 48.61% to 73.48% | 56.82 | 75.30% | 91.00% |
| > 4.440 | 90 | 69.90% to 98.22% | 63.6 | 50.42% to 75.07% | 53.64 | 88.20% | 67.70% |
| > 6.105 | 55 | 32.21% to 74.18% | 90.9 | 80.42% to 96.05% | 45.91 | 88.10% | 62.30% |

Figure 5 Legend (Supplementary Data): ROX distribution by outcome at 24h with cutpoints

