## Supplementary figures and images for "Evaluation of the ROX index in SARS-CoV-2 Acute Respiratory failure treated with both High-Flow Nasal Oxygen (HFNO) and Continuous Positive Airway Pressure (CPAP)"

### Figure 5 Supplementary data

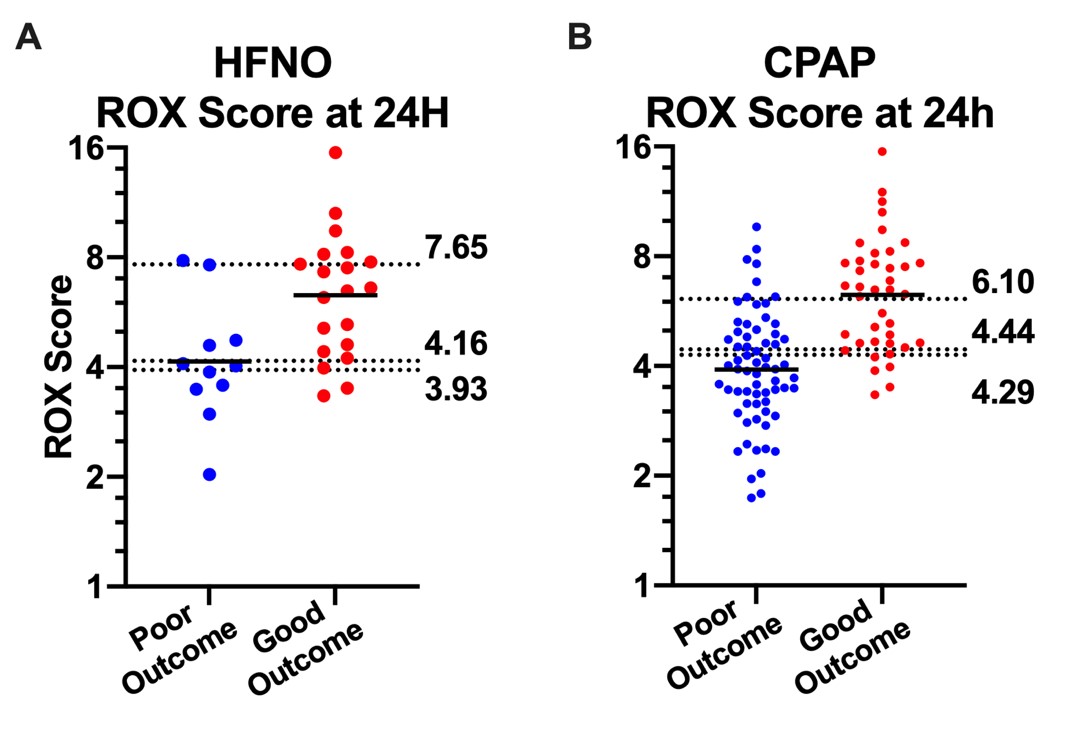
